# Early multidrug treatment of SARS-CoV-2 (COVID-19) and decreased case fatality rates in Honduras

**DOI:** 10.1101/2021.07.21.21260223

**Authors:** Sidney Ontai, Li Zeng, Miguel Sierra Hoffman, Fernando Valerio Pascua, Vincent VanBuren, Peter A McCullough

## Abstract

**INTRODUCTION:** Within 2 months of first detection of SARS-CoV-2 in Honduras, its government promoted nationwide implementation of multi-drug COVID-19 inpatient and outpatient treatment protocols. This was associated with a case fatality rate decrease from 9.33% to 2.97%. No decrease was seen in Mexico, a similar Latin American country that did not introduce multi-drug treatment protocols at that time.

**OBJECTIVE:** The primary objective of the study was to use statistical process control to assess the likelihood that the decrease in case fatality rate in Honduras was due to chance, using Mexico as a control country.

**METHODS:** Fourteen day running average COVID-19 case fatality rates in Honduras and Mexico were used to create Shewhart control charts during the first 6 months of the epidemic. The date of implementation in Honduras of the inpatient and outpatient multi-drug COVID-19 protocols were plotted on control charts, with a Mexican COVID-19 case fatality control chart for comparison.

**RESULTS:** The case fatality rate for COVID-19 in Honduras dropped below the lower control limit 9 days after implementation of an inpatient and outpatient multi-drug therapeutic protocol, from an average 9.33% case fatality rate to 5.01%. The Honduran COVID-19 case fatality rate again dropped below the lower control limit to 2.97%, 17 days after launching a substantial government program to make the protocol medications accessible to underserved areas. Shewhart control chart plots of case fatality rates in Honduras suggest a plausible temporal association between the implementation dates of both the initial protocol implementation on May 3, 2020, and the outreach effort on June 10, 2020, and statistically significant control chart anomalies. No control chart anomalies were seen during that time in Mexico.

**CONCLUSION:** Decreases in COVID-19 case fatality rates in Honduras were associated with both the initial publication a multi-drug COVID-19 therapeutic protocol and a subsequent outreach program.

## BACKGROUND

Honduras is a low middle-income country with 66 percent of its population in poverty in 2016, and with 20 percent of Hondurans living in extreme poverty—that is, on less than US$1.90 per day.^1^ Honduras experienced its first case of COVID-19 documented by SARS CoV-2 RT-PCR testing on March 10, 2020.^2^

As the COVID-19 pandemic unfolded in New York in the spring of 2020, Honduras faced the potential collapse of its health care system from COVID-19, with but 19 critical care physicians and 125 intensive care unit (ICU) beds to serve a population of 9.9 million.^3^ By contrast, New York City with a population of 8.9 million, had 1060 critical care physicians and 1644 ICU beds.^4^ Still recovering from a recent Dengue epidemic, the Honduran government responded proactively to the COVID-19 pandemic based on the recommendations of its critical care and infectious disease consultants. Honduran physicians theorized that a multi-mechanism approach (MMA) to treating COVID-19 could block the inflammation, immune system disruption, and hypercoagulation that initial reports suggested as the cause for mortality from SARS CoV-2 infections. They created the MMA protocol based on the hypothesis that COVID-19 mortality resulted less from direct viral infection and more from the self-perpetuating cytokine storm and thrombosis triggered by the virus that, once triggered, increased independently of viral replication (Figure 1). Given Honduras’ limited resources, the MMA protocol used repurposed, inexpensive medications already proven safe and effective for other indications, for which *in vitro* and clinical evidence suggested clinical efficacy against COVID-19. The MMA protocol also optimized oxygenation with high-flow O_2_ therapy and self-pronation rather than mechanical ventilation whenever possible, conserving limited intensive care resources. In mid-April 2020, Honduran physicians began treating COVID-19 patients with the MMA protocol immediately upon admission to the hospital, as well as in outpatient clinics immediately on diagnosis with COVID-19. Decreased morbidity and mortality were observed in patients receiving the MMA inpatient protocol, documented in a peer-reviewed retrospective cohort study that showed a decrease ICU length of stay by 5.4 days with a trend towards decreased mortality.^5^

**FIGURE 1:**
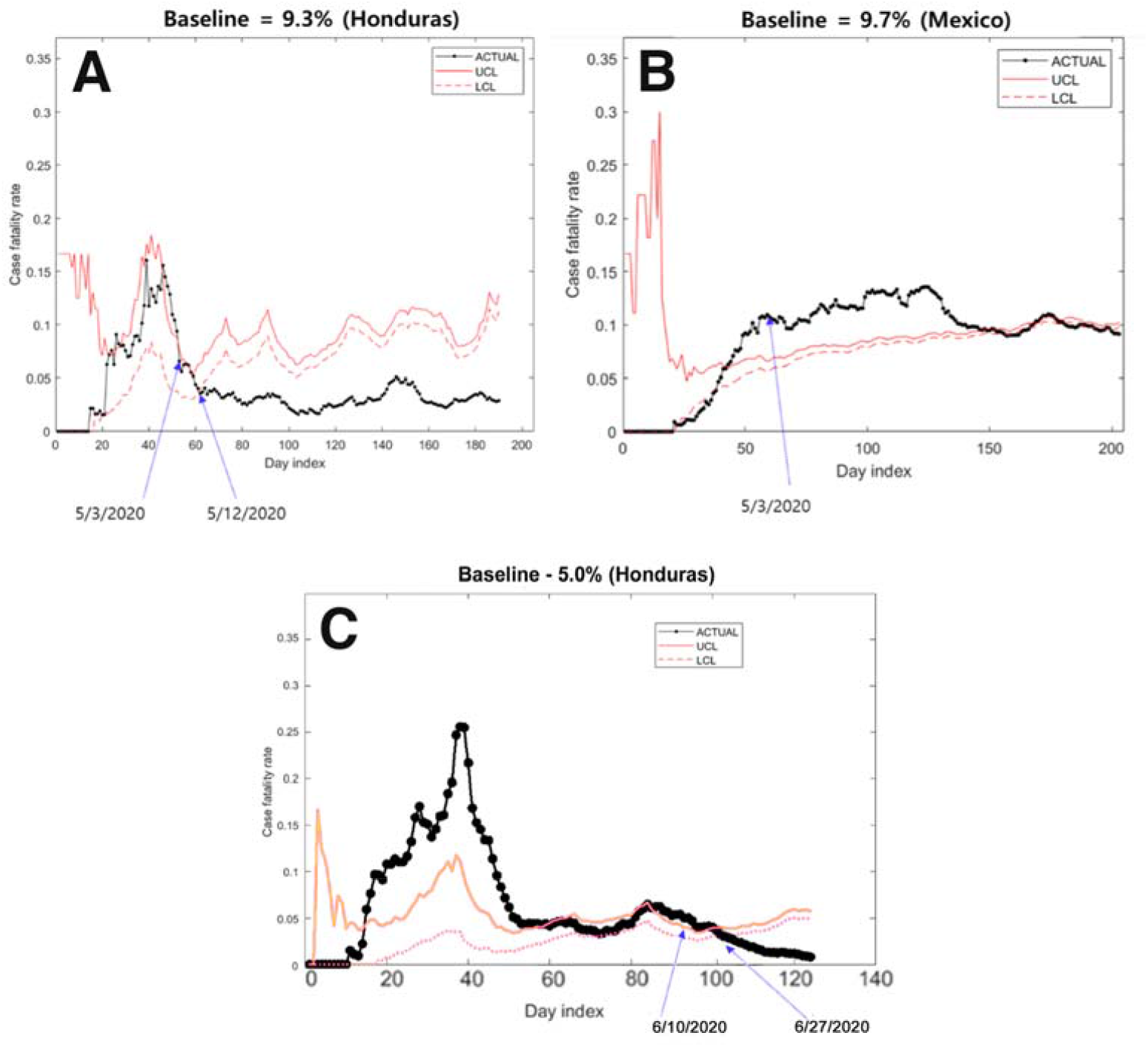
Shewhart control chart upper and lower control limits for 14 day rolling average case fatality rate. Control limits were calculated using a baseline which was the average case fatality as of May 3, 2020 for Honduras (A) and Mexico (B); and as of June 10, 2020 for Honduras (C).

The MMA protocol^6^ (Table 1A) medications have yet to be labeled by the FDA as effective for COVID-19 therapy, although they have been labeled as safe for non-COVID-19 indications. After the initial promising results, the protocol was promoted by the Honduran Health Department on May 3, 2020, in a nationally televised educational program for health care professionals, detailing recommendations on treating COVID-19 with MMA for inpatients and outpatients. The Honduran Health Department hosted additional Zoom™ video meetings to educate clinicians throughout the month of May 2020. The inpatient MMA protocol was marketed as “CATRACHO,” an acronym of the protocol’s components that references General Florencio Xatruch, the Honduran leader who defeated aggressors from the U.S. in 1856. Hondurans refer to themselves as “catrachos,” a Nicaraguan pronunciation of the Catalan surname Xatruch.^7^

**TABLE 1:**
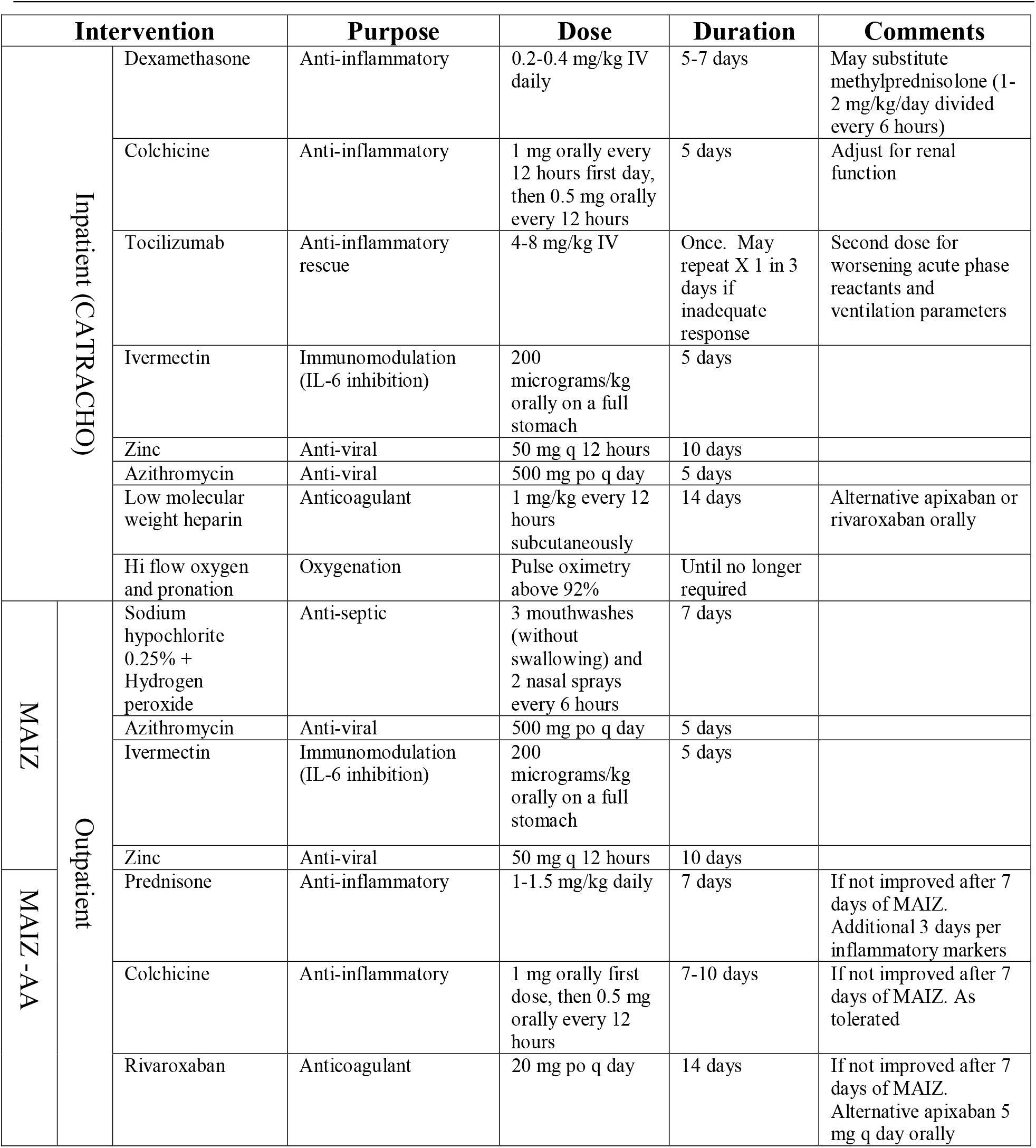
Inpatient and Outpatient Medication Protocols for Honduras’ Multi-Mechanism Approach to COVID-19 Therapy

As the pandemic in Honduras spread, the number of COVID-19 outpatients overwhelmed existing clinic capacity. The Honduran government elected to create “medical brigades” that performed home visits, identifying underserved COVID-19 patients early and immediately dispensing outpatient protocol medications. The initial 7-day course of outpatient therapy was prepackaged as “MAIZ” (Spanish for “corn,” Honduras’ staple food), an acronym for mouthwash of sodium hypochlorite and hydrogen peroxide, azithromycin, ivermectin, and zinc. Patients that continued to have COVID-19 symptoms after 7 days were prescribed an enhanced outpatient therapy, “MAIZ-AA”, which added anti-inflammatory medications (colchicine and prednisone) and an anticoagulant (rivaroxaban).

Within three months after the initiation of the MMA protocols in Honduras, clinicians worldwide began to publish reports supporting the efficacy against SARS CoV2 infections of the main components of the MMA protocols: namely colchicine,^8 9^ ivermectin,^10^ tocilizumab,^11^ dexamethasone,^12^ and full dose heparin.^13^

## METHODS

Due to the urgency of the pandemic and the paucity of local healthcare resources, it was impractical to prospectively organize randomized, controlled, double-blinded trials to evaluate the MMA protocol’s efficacy against COVID-19 prior to its implementation in Honduras. An alternative methodology, statistical process control, is a well validated approach initially developed to improve manufacturing outcomes at the Bell Laboratories about a century ago. Statistical process control has gained increasing acceptance in health care applications.^14^ It is less resource intensive, achieving statistical significance through frequent measurements over time rather than by large patient sample sizes, and represents results in easy to comprehend control charts.^15^

Shewhart control charts were used to compare COVID-19 case fatality rates in Honduras (Figure 2A) with those in a control country, Mexico (Figure 2B), which has a similar climate, and whose population has comparable age, demographics, socio-economic status, and experienced nearly identical case fatality rate increases during its initial exposure to SARS CoV2. The publicly available World Health Organization online dashboard^16^ provided Honduran and Mexican case fatality data.

## RESULTS

On May 3, 2020, the Honduran Department of Health encouraged physicians to implement the MMA protocol (Table 1) for inpatients (CATRACHO protocol) immediately on hospital admission, and for outpatients (MAIZ protocol) immediately on diagnosis. If outpatients continued to have symptoms after 7 days, the “MAIZ-AA” anti-inflammatory and anticoagulant medications (colchicine, prednisone and rivaroxiban) were added. The Honduran COVID-19 case fatality rate dropped below the Shewhart control chart’s lower control limit on May 12, 2021 (Figure 2A), nine days after the publication of the MMA recommendations. The baseline case fatality rate of 9.3% was used to calculate the control limits was the average case fatality rate in Honduras on May 3, when the MMA inpatient and outpatient protocols were first implemented. Synchrony exists between the case fatality rate control chart anomaly in Honduras and the dates on which its government published the MMA COVID-19 treatment protocol.

Figure 2C shows that in Honduras, the case fatality rate dropped below the Shewhart control limit a second time 17 days after initiation of the government rural outreach program to distribute outpatient protocol medications (MAIZ) (Table 1) on June 10, 2020. By contrast, the Mexico data (Figure 2 B) did not show any dropping trends below case fatality rate lower control limits during May or June 2020.

## DISCUSSION

The initial implementation of the MMA protocols was associated with a 6.36% decrease in COVID-19 case fatality rate in Honduras, from 9.33% before May 3, 2020 to 2.97% after. This suggests the number needed to treat (NNT) is 16 patients to prevent one COVID-19 fatality for the combined inpatient (CATRACHO) and outpatient (MAIZ/MAIZ-AA). Recalculating the control chart limits using the 5.01% average case fatality rate on June 10, 2020 as baseline demonstrated a statistically significant drop below the lower control limit on June 27, 2020. This case fatality rate decrease from 5.01% to 2.97% suggests the avoidance of 1 COVID-19 fatality for every 49 outpatients treated by the additional outreach initiative by Honduran medical brigades implementing the MAIZ/MAIZ-AA outpatient therapeutic protocol.

Our findings demonstrate the utility of statistical process control methodology for quickly and efficiently evaluating and monitoring the efficacy of therapies for COVID-19, which could be generalizable to other emergent conditions.

## Data Availability

All COVID-19 case fatality data were obtained from the publicly available World Health Organization Coronavirus Disease (COVID-19) Dashboard.

https://covid19.who.int/table

